# Statistical Relationship Between Bitcoin and Synthetic Opioid Mortalities: Are DEA Enforcement Actions Aligning with Trends in Drug Related Deaths?

**DOI:** 10.1101/2024.03.27.24304987

**Authors:** Daihun Kang

## Abstract

**Background:** The intersection of cryptocurrency, especially Bitcoin, with public health issues, particularly synthetic opioid-related deaths, presents an emerging field of study. This research explores the statistical relationship between Bitcoin market fluctuations and synthetic opioid mortalities, against the backdrop of Drug Enforcement Administration (DEA) enforcement actions.

**Methods:** Utilizing data from 2009 to 2022, this study employs Pearson correlation and linear regression models to investigate the relationship between annual Bitcoin price fluctuations and synthetic opioid-related death rates, alongside DEA domestic arrest trends.

**Results:** A pronounced positive correlation (r = 0.92) was found between Bitcoin price changes and synthetic opioid mortality rates, with the Bitcoin price accounting for approximately 84.78% of the variance in opioid-related deaths (R² = 0.8478). The analysis also notes a disparity between increasing synthetic opioid fatalities and decreasing DEA domestic arrests.

**Conclusions:** The findings reveal significant correlations between Bitcoin price fluctuations and synthetic opioid-related fatalities, highlighting potential gaps in current drug enforcement strategies. This study underscores the need for an interdisciplinary approach to address the complexities introduced by cryptocurrency in the opioid crisis and suggests the necessity of integrating financial and public health strategies to combat emerging drug abuse trends.

## Introduction

The burgeoning adoption of Bitcoin and other cryptocurrencies has significantly impacted various sectors globally, prompting extensive research into their economic and social repercussions [1]. The volatile nature of cryptocurrency markets, especially Bitcoin, has been a topic of considerable interest and concern among economists, investors, and policymakers [2]. This interest is heightened by the parallel increase in synthetic opioid-related incidents, which has become a pressing public health crisis, particularly in states like California [3]. Reports of individuals experiencing severe physical reactions, akin to a state of paralysis often described colloquially as “zombification,” after consuming potent synthetic opioids like fentanyl, are becoming alarmingly common [4]. Additionally, there have been instances where law enforcement officers, merely exposed to the drug’s airborne particles, have become incapacitated, underscoring the drug’s potent nature [5].

The intersection of these two seemingly disparate trends – the rise of cryptocurrencies and the opioid epidemic – raises intriguing questions about the potential connections between the digital economy and public health crises. Anecdotal evidence and portrayals in media, such as television dramas and films, frequently suggest that drug cartels are increasingly turning to Bitcoin and other digital currencies as a means to launder money [6]. These narratives depict a shift from traditional money laundering methods to opaquer, decentralized financial networks, facilitated by cryptocurrencies. This shift is believed to not only complicate the efforts to combat drug trafficking but also potentially impact the dynamics of drug abuse and associated mortality rates.

Prompted by these observations, this study hypothesizes that there may be a discernible relationship between the surge in synthetic opioid usage, particularly in the United States, and fluctuations in Bitcoin prices. This hypothesis stems from the presumption that as synthetic opioids become more prevalent, the resultant increase in illicit drug trade could be mirrored in the dynamics of Bitcoin transactions and value. Therefore, this research aims to explore the potential correlation between synthetic opioid-induced mortality rates and Bitcoin price fluctuations, providing insights into how emerging digital financial markets might be inadvertently influencing public health trends, especially concerning the ongoing opioid epidemic. Through this investigation, we seek to offer a comprehensive understanding of these complex interdependencies, contributing to the broader discourse on the socioeconomic impacts of modern technologies and public health challenges.

## Materials and Methods

This study utilized data from the years 2009 to 2022, focusing on annual Bitcoin prices and synthetic opioid-related mortality rates. The initial exploratory data analysis involved Pearson’s correlation to assess the relationship between these two factors. Subsequent analyses included linear regression to determine the impact of Bitcoin price fluctuations on synthetic opioid-related mortality rates. These statistical analyses were conducted using the Python programming language, employing libraries such as Pandas for data management and SciKit-Learn for regression analysis. This approach allowed for a detailed examination of the dynamic interactions between the variables under study.

Data sources included Bitcoin price information obtained from CoinCodex [7] and synthetic opioid-related death counts provided by the National Center for Health Statistics [8]. Additionally, U.S. Gross Domestic Product (GDP) figures were sourced from the U.S. Bureau of Economic Analysis, retrieved via the Federal Reserve Bank of St. Louis database [9]. The data on domestic arrests was retrieved from the United States Drug Enforcement Administration (DEA) website [10].

This study utilized publicly available data, which did not contain any personally identifiable information, ensuring that specific individuals could not be identified. Therefore, obtaining informed consent was not applicable. The analysis focused on annual Bitcoin prices and synthetic opioid-related mortality rates from 2009 to 2022. The data, including Bitcoin prices from CoinCodex and mortality rates from the National Center for Health Statistics, as well as U.S. GDP figures and DEA domestic arrests data, are all publicly accessible and do not involve private individual information.

## Results

This study meticulously analyzed a comprehensive dataset, detailed in Table 1, which presents annual data from 2009 to 2022 on various indicators related to drug abuse. These indicators include drug abuse deaths, opioid-related fatalities, and deaths due to synthetic opioids other than Methadone. Additionally, the dataset encompasses associated metrics such as cocaine and benzodiazepine fatalities, DEA domestic arrests, U.S. population figures, and GDP trends.

**Table 1.**
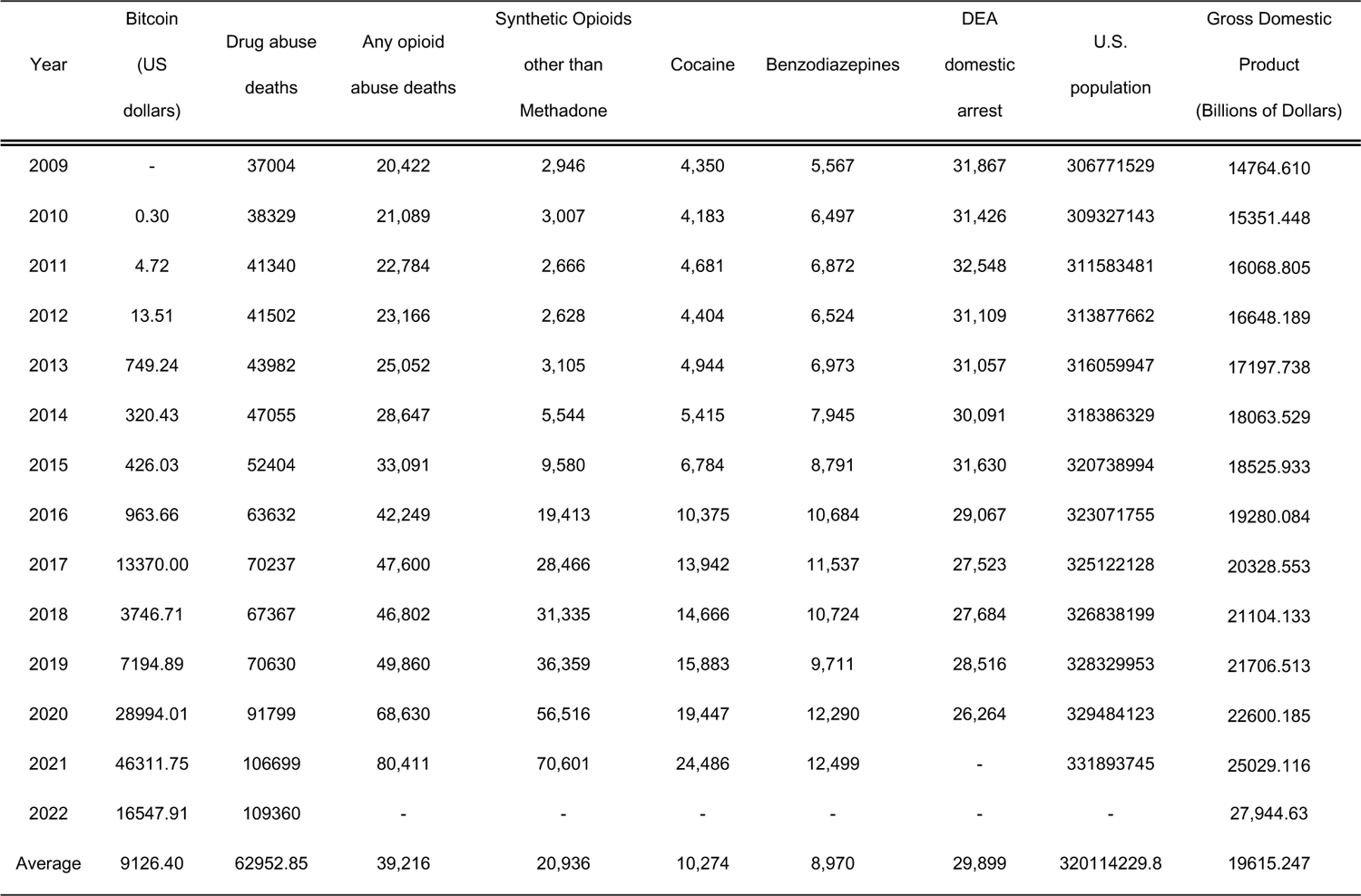
Annual Trends in Bitcoin Valuation, Drug-Related Deaths, and Drug Enforcement Administration (DEA) Arrests (2009-2022)

The analysis identified a significant escalation in drug abuse fatalities, particularly those attributed to synthetic opioids, juxtaposed against the backdrop of fluctuating GDP and demographic transitions (Fig 1). Despite the volatility observed in the Bitcoin market, with marked declines following substantial hikes, a notable rise from approximately $0.30 in 2010 to $46,311.75 in 2021 was documented (Fig 2) [11].

**Figure 1.**
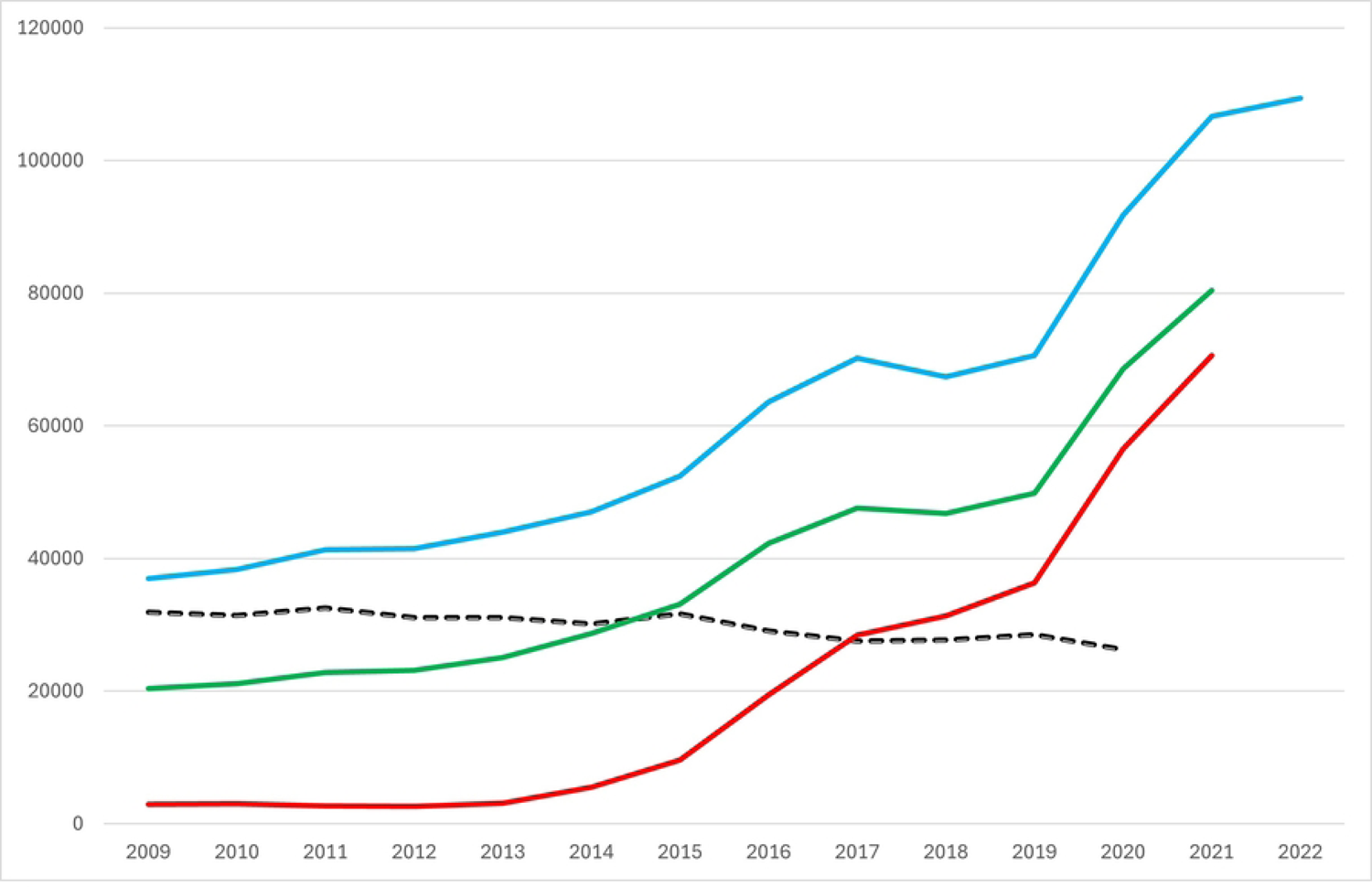
Synthetic Opioids other than Methadone.

**Figure 2.**
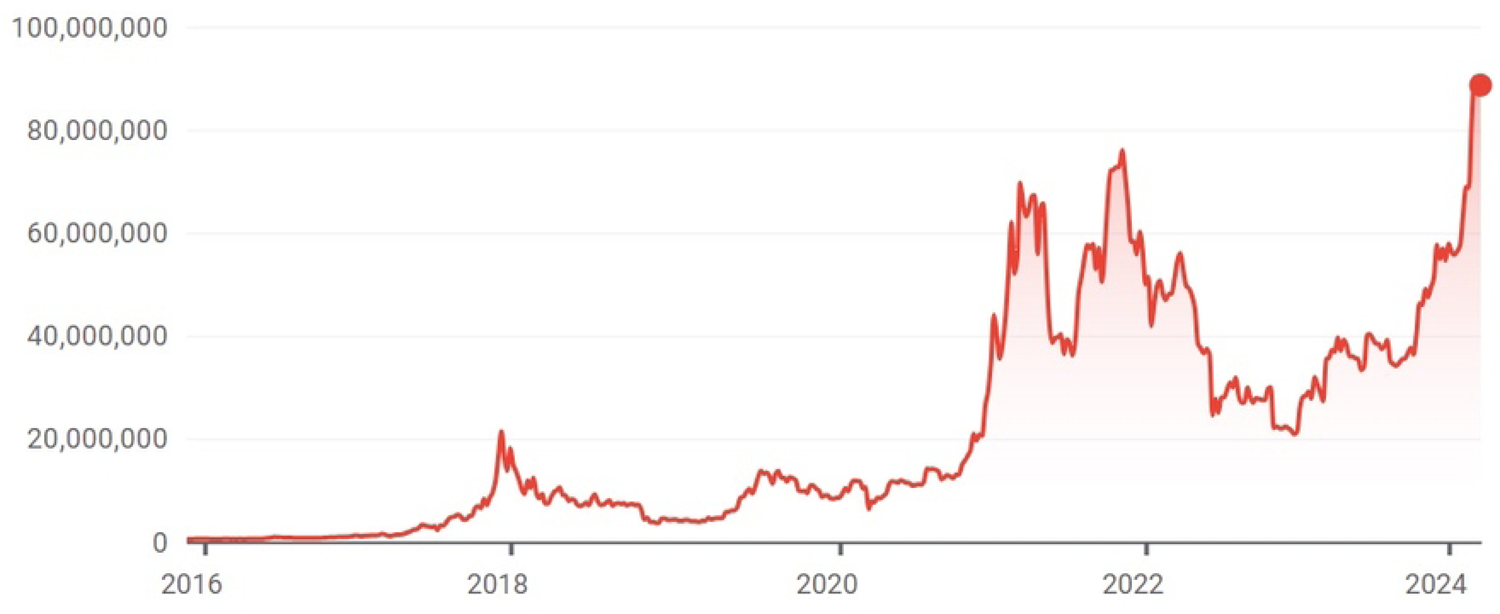
Bitcoin Price Fluctuations (2016-2024) in US Dollars

Blue: Total drug abuse deaths. Green: Deaths from any opioid use. Red: Deaths from synthetic opioids, excluding Methadone. Black dashed: Drug Enforcement Administration domestic arrests.

A closer examination, as depicted in Figure 3, reveals a synchronicity between Bitcoin price fluctuations and synthetic opioid-related deaths, with synthetic opioid fatalities consistently rising year-over-year, despite the Bitcoin price experiencing significant downturns in 2017 and 2021. Conversely, a continuous decline was observed in DEA domestic arrests, suggesting a possible disconnection with the prevailing drug abuse trends.

**Figure 3.**
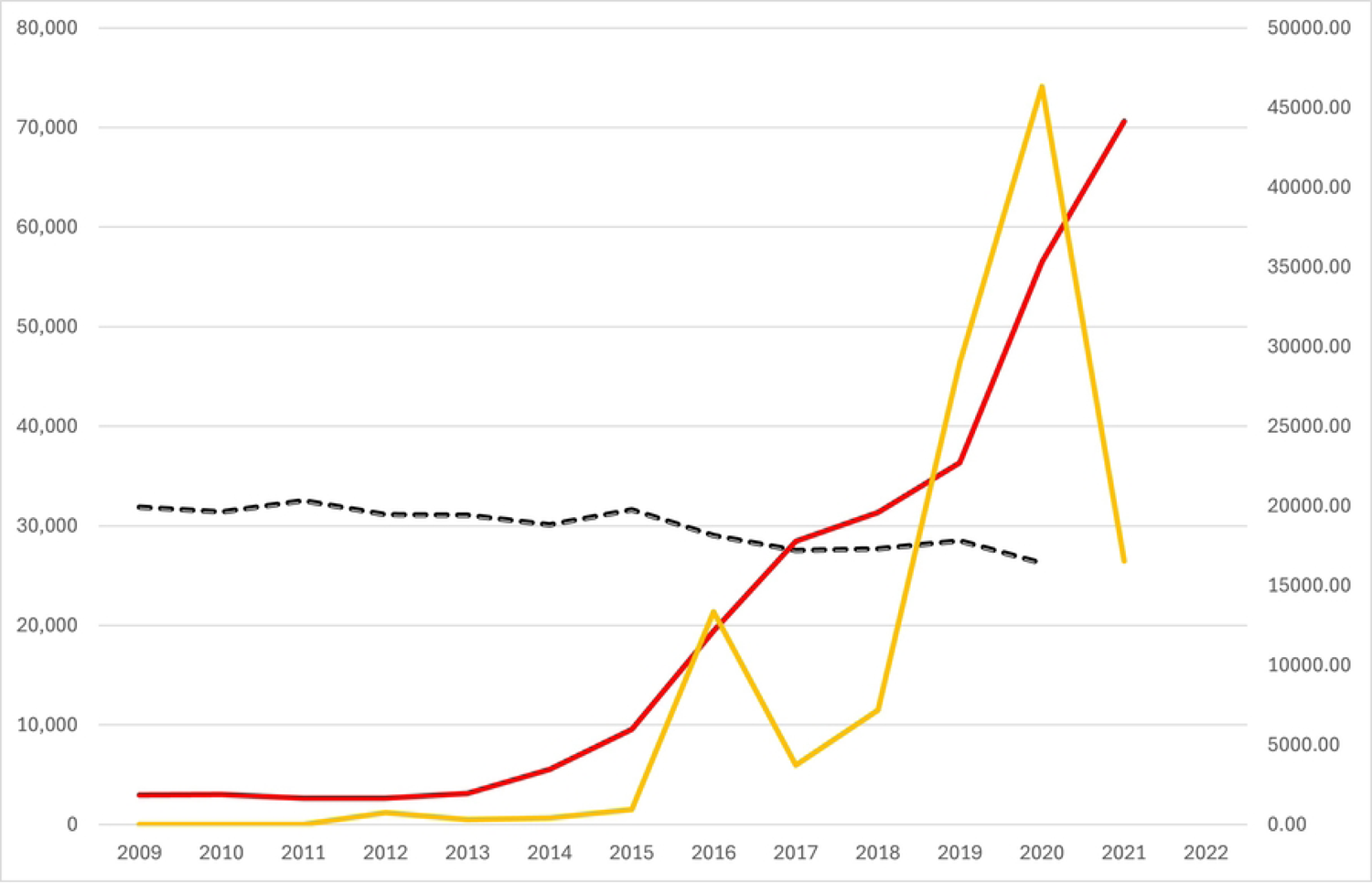
Trends in Bitcoin Price, Synthetic Opioid-Related Deaths, and Drug Enforcement Administration (DEA) Domestic Arrests (2009-2022)

Yellow line: Bitcoin Price in US Dollars.

Red line: Number of deaths due to Synthetic Opioid Use. Black dashed line: Number of DEA Domestic Arrests.

Employing Pearson’s correlation analysis, the statistical examination unveiled a pronounced positive correlation (r = 0.92) between the volatility in Bitcoin prices and the incidence of synthetic opioid-related deaths. This strong correlation suggests a significant linkage; rising Bitcoin valuations seem to be paralleled by increases in synthetic opioid-related fatalities.

Delving deeper through linear regression analysis, we obtained the following results:

1. Bitcoin Price as an Independent Variable: The derived regression equation was: Synthetic Opioid Deaths = 1.4406 × Bitcoin Price + 10,178.61. Herein, a unit increase in the Bitcoin price corresponds to a 1.4406 increase in the count of synthetic opioid-related deaths. This model elucidates approximately 84.78% of the variance (R² = 0.8478), indicating a substantial proportion of synthetic opioid deaths can be accounted for by fluctuations in Bitcoin price.
2. Synthetic Opioid Deaths as an Independent Variable: The corresponding regression equation manifested as: Bitcoin Price = 0.5885 × Synthetic Opioid Deaths − 4,694.75.

This denotes that each increment in synthetic opioid-related fatalities is associated with an average upsurge of 0.5885 in Bitcoin valuation, reaffirming the model’s substantial explanatory capacity (R² = 0.8478).

The analysis highlights significant correlations between economic indices and public health outcomes. It is imperative, however, to underscore that while the correlations and regression models delineate clear patterns, they do not, in isolation, confirm causation. This calls for an advanced investigation to decipher the causative dynamics underpinning these relationships.

Critically, an examination of the temporal trends reveals that notwithstanding the substantial swell in drug abuse and synthetic opioid-related fatalities, DEA domestic arrests have not paralleled this trend. Specifically, while the average annual count of DEA domestic arrests stood at approximately 29,899, there was a conspicuous downturn to 26,264 arrests by the year 2020. This decline in drug enforcement arrests is starkly at odds with the surging drug abuse-related mortality rates. Such discordance underscores potential disparities or evolutions in drug enforcement strategies, raising pivotal questions regarding the efficacy and direction of existing drug policy frameworks against the backdrop of an escalating opioid crisis.

## Discussion

### Main Finding of This Study

The pivotal discovery of our investigation is the substantial positive correlation between fluctuations in the Bitcoin market and the rates of synthetic opioid-related fatalities. This connection, quantified by a correlation coefficient of r = 0.92, demonstrates a significant association that suggests economic events within the realm of digital currencies may have tangible impacts on public health crises, specifically the opioid epidemic. This finding underscores the complex interplay between financial markets and drug abuse trends, challenging traditional perceptions of their separate domains.

### What is Already Known on This Topic

Prior research has hinted at the potential intersections between cryptocurrency and illegal drug markets, primarily focusing on the anonymity and lack of regulation within digital currency systems that can facilitate illicit trade [12]. Studies have also explored the rise in opioid-related deaths, emphasizing the growing concern around synthetic opioids such as fentanyl [13]. However, there has been a gap in directly linking the economic behaviors of cryptocurrency, like Bitcoin, with tangible public health outcomes, particularly in the realm of drug-related fatalities [14].

### What This Study Adds

This research marks a significant advancement in the field by meticulously establishing an empirical linkage between the fluctuations of the Bitcoin market and the escalating incidences of synthetic opioid-related fatalities. Prior to this analysis, the academic and medical communities have primarily focused on either the rise of digital currency or the opioid epidemic as isolated phenomena. By bridging these two crucial areas, our study unveils the unforeseen impacts of cryptocurrency market dynamics on public health, particularly within the sphere of synthetic opioids.

Our findings extend beyond the traditional narrative, shedding light on how economic trends, especially in emerging markets like cryptocurrencies, can have profound and tangible effects on societal health crises. This integration of economic and public health data provides a novel perspective, suggesting that the volatility inherent in digital financial markets could serve as a barometer for predicting trends in drug-related health issues. This insight is pivotal for public health officials and policymakers, who traditionally may not have considered financial market behaviors when devising strategies to combat the opioid crisis.

Moreover, this study adds a critical dimension to the understanding of drug enforcement effectiveness. By highlighting the discrepancy between the rise in opioid fatalities and the decrease in DEA domestic arrests, it prompts a reevaluation of current strategies employed in the war against drugs. This divergence may suggest that conventional methods and resources are becoming increasingly inadequate in addressing the nuances of modern drug trafficking, which may be evolving in part due to the advent of cryptocurrencies. Consequently, our research underscores the urgent need for innovative approaches and tools in drug enforcement and policy-making that are adaptive to the changing landscape facilitated by digital currencies.

In addition, our work introduces a pivotal discussion regarding the regulatory implications for cryptocurrencies. While digital currencies have been celebrated for their innovation and democratizing potential, our study underscores the dark side of this technological advancement, particularly its utilization in illicit drug markets. By empirically demonstrating the correlation between Bitcoin market fluctuations and opioid-related deaths, this research underlines the necessity for comprehensive regulatory frameworks that balance the benefits of cryptocurrencies with the imperative to prevent their misuse in activities detrimental to public health.

Furthermore, this research acts as a call to action, not only for policymakers and law enforcement agencies but also for the cryptocurrency community itself. It challenges stakeholders across sectors to acknowledge and address the unintended consequences of digital financial innovations on societal well-being. In doing so, it lays the groundwork for a multidisciplinary approach to tackling the opioid epidemic, one that incorporates financial regulation, public health strategy, and law enforcement efficiency.

Lastly, this study enriches the academic discourse by offering a foundation for future research in this emergent interdisciplinary field. The significant correlation identified here opens numerous avenues for further exploration, from the mechanisms underlying the relationship between cryptocurrency and opioid fatalities to the impact of specific regulatory interventions. As such, this research not only contributes to our current understanding but also sets the stage for a new era of inquiry that seeks to unravel the complex interdependencies between our digital economic systems and public health outcomes.

## Conclusion

This study enriches the discourse on the intersection of digital finance and public health by providing empirical evidence and a new analytical lens through which to view the opioid epidemic. It calls for a concerted effort among stakeholders to forge comprehensive strategies that address the multifaceted challenges presented by the integration of cryptocurrency markets and public health concerns. Future research should build on these findings, exploring the causal pathways and developing targeted interventions that can mitigate the adverse effects of this intersection on society.

## Data Availability

The data available from the provided sources include: U.S. Gross Domestic Product (GDP) historical data from the Federal Reserve Bank of St. Louis. Historical Bitcoin price data from CoinCodex. Statistics on domestic arrests from the U.S. Drug Enforcement Administration (DEA). Bitcoin price history and current prices from Google Finance. These resources offer publicly accessible data relevant for economic, legal, and financial research. For full details, visit the respective websites: https://fred.stlouisfed.org/series/GDP https://coincodex.com/article/31832/bitcoin-price-history/. https://www.dea.gov/data-and-statistics/domestic-arrests. https://www.google.com/finance/quote/BTC-KRW?sa=X&ved=2ahUKEwiksrKD-_qEAxU8h1YBHT_mBvkQ-fUHegQIJhAf&window=MAX.

## Acknowledgments

None.

